# DNA Methylation as a Mediator of Cardiovascular Disease Risk in Relation to PTSD Severity: Identification of Potential Epigenetic Biomarkers

**DOI:** 10.64898/2026.01.22.26344545

**Authors:** Md Mainul Hasan Sarker, Andrew Ratanatharathorn, Jan Dahrendorff, Chengqi Wang, Agaz H. Wani, Allison E. Aiello, Annie Qu, Karestan C. Koenen, Alicia K. Smith, Derek E. Wildman, Monica Uddin

## Abstract

Post-traumatic stress disorder (PTSD) is associated with increased cardiovascular disease (CVD) risk, yet the epigenetic mechanisms underlying this link remain unclear. We investigated whether DNA methylation (DNAm) within Conserved Regions of Systemic Interindividual Variation (CoRSIVs), genomic regions showing stable within-individual, but variable between-individual methylation, mediates the association between PTSD symptom severity (PTSS) and CVD.

We analyzed blood-derived DNAm from three cohorts (DNHS: discovery; GTP and NHS: replication), focusing on 7,694 CoRSIV CpGs profiled with the Illumina MethylationEPIC BeadChip. Logistic regression related CpGs to PTSS and CVD, adjusting for demographic and trauma-related covariates. CpGs nominally associated (p<0.05) with both PTSS and CVD in DNHS were then tested using causal mediation analysis.

In DNHS, 27 CpGs were nominally associated with both PTSS and CVD, with seven showing nominal mediation (p<0.05). Across cohorts, six of these seven displayed mediation effects in a consistent direction in at least one replication cohort, and three CpGs showed concordant mediation directions across all three cohorts. Notably, cg07941916 (C5orf56/IRF1-AS1) and cg20545458 (intergenic) exhibited positive mediation in DNHS with the same direction in GTP, implying that higher PTSS is associated with methylation changes that correspond to higher CVD risk, whereas all mediation effects in NHS were negative, consistent with its healthier, lower-risk profile. These loci map to immune and inflammatory pathways, alongside other mediators annotated to neuronal/stress-aging and autonomic processes.

Overall, DNAm variation within CoRSIVs may partially mediate PTSD-related CVD risk and nominates specific CpGs as hypothesis-generating epigenetic biomarkers that require validation in larger, ancestrally diverse longitudinal cohorts.

## 1. Introduction

### Background

Cardiovascular disease (CVD) remains the leading cause of global mortality, responsible for almost 20.5 million deaths annually, and approximately one-third of all casualties worldwide (1–3). CVD resulted in 941,652 deaths in the United States in 2022 (4,5). In addition to its impact on mortality, CVD in the United States yields annual costs exceeding $400 billion, which includes direct costs of $233.3 billion and indirect costs of $184.6 billion from 2020 to 2021, affecting both mortality and productivity (5,6). The global incidence of CVD is increasing despite advancements in treatment and prevention (7): ischemic heart disease is the primary contributor to disability-adjusted life years (DALYs) globally (2). This ongoing challenge underscores the importance of identifying new risk factors and establishing preventive measures to reduce the incidence of CVD.

Extensive research has established classic CVD risk factors such as hypertension, hyperlipidemia, smoking, diabetes, and obesity, yet these do not fully account for observed CVD events. Increasing attention has therefore been directed toward psychosocial stressors (8). Traumatic stress and conditions such as post-traumatic stress disorder (PTSD) can induce heightened inflammation and autonomic dysregulation, and depression, anxiety, and chronic stress predict cardiovascular outcomes with effect sizes comparable to smoking and obesity (8,9). Psychological stress and its potential consequences, particularly PTSD, have been identified as significant factors in the risk of CVD (10). PTSD, characterized by intrusive memories, avoidance, negative mood or cognitions, and hyperarousal after trauma exposure (11), is common given the ubiquity of trauma (12,13), and numerous studies link PTSD to elevated CVD morbidity and mortality (10,14–16). PTSD is now recognized as a systemic condition involving coordinated changes in central nervous, immune/inflammatory, and neuroendocrine–metabolic systems, providing plausible biological pathways that could connect traumatic stress to adverse cardiovascular outcomes (17–19).

Despite the growing evidence linking PTSD and CVD, the specific biological mechanisms underlying this association remain poorly understood. Genetic studies have identified shared etiological factors between PTSD and hypertension, including chronic inflammation (CRP, IL6), immune dysfunction, metabolic traits (waist-to-hip ratio), insomnia, and substance use (20); whereas coronary artery disease, heart failure, and atrial fibrillation appear to be primarily distinguished by electrophysiological etiology and do not exhibit a significant genome-wide genetic correlation with PTSD (21), In contrast, the role of epigenetic modifications, particularly DNA methylation (DNAm), in mediating the PTSD-CVD relationship remains limited and inconclusive (22). Studies investigating DNAm in the context of PTSD and stress-related disorders have shown that DNAm differences affect immunological, endocrine, and vascular pathways, which are vital to the pathogenesis of CVD (23–25). Differential DNAm patterns may assist in diagnosing and stratifying molecular malfunctions before clinical symptoms appear. Blood-based DNAm, particularly within CoRSIVs regions, may be especially informative for studying systemic disease processes. CoRSIVs are specific genomic regions that exhibit highly consistent DNAm patterns across multiple tissues within an individual (spanning ectodermal, mesodermal, and endodermal lineages) while showing substantial inter-individual variability (26). While the goal of EWAS is often to identify purely epigenetic variants (metastable epialleles), prior analyses indicate that genetic influence on CoRSIV methylation is extremely strong, with approximately 60% of interindividual variation explained by cis-acting genetic variation (27). Crucially, regardless of the underlying etiology, this cross-tissue consistency means that methylation profiles measured in easily accessible tissues such as peripheral blood can provide a reliable indication of epigenetic regulation in internal organs. This is supported by findings that methylation in blood correlates with methylation and gene expression in internal organs, allowing researchers to study epigenetic states in less accessible but disease-relevant tissues, including those involved in cardiovascular pathology (27).

The present study seeks to identify DNAm-based epigenetic biomarkers that may mediate the link between PTSD severity and CVD risk, with a specific focus on DNAm patterns within CoRSIVs. We hypothesize that specific DNAm signatures will mediate the relationship between PTSD symptom severity and cardiovascular disease risk, and that these epigenetic changes will be enriched in biological pathways relevant to both stress response and cardiovascular function. Using the Detroit Neighborhood Health Study (DNHS) as a discovery cohort and replicating findings in the Nurses’ Health Study (NHS) and Grady Trauma Project (GTP), we identified CpG sites associated with both PTSD symptoms and CVD risk. We further investigated whether these DNAm patterns mediate the PTSD–CVD relationship and explored the cellular pathways associated with these relevant CpGs. This initial investigation reveals nominally significant patterns that require replication in larger cohorts. These findings suggest potential avenues for identifying trauma-exposed individuals who may face elevated cardiovascular disease risk, though further validation in larger cohorts is needed before clinical application. Our work represents an important first step toward developing biomarker-based approaches for personalized risk assessment in trauma survivors.

## 2. Materials and Methods

### Participants and Study Design

We analyzed data from three independent cohorts to investigate the association among post-traumatic stress disorder severity (PTSS), DNAm, and cardiovascular disease (CVD). These included a discovery cohort from the Detroit Neighborhood Health Study (DNHS), and two replication cohorts: Nurses’ Health Study (NHS), and the Grady Trauma Project (GTP). Each study was approved by the relevant institutional review board, and all participants provided written informed consent and details about each cohort are given in the supplementary file.

### DNA methylation: Quantifying and processing

Methylation profiling of whole blood was measured using the Illumina MethylationEPIC BeadChip. The study applied an identical preprocessing and QC pipeline to each cohort (28). Raw intensity data were processed in R (v4.4.2), excluding any sample with a probe detection call rate below 90% or with an average fluorescence intensity under 50% of the experiment-wide mean or below 2000 AU. Probes with detection p-values > 0.01 were marked missing, and any probe absent in >10% of samples was removed (29). In addition cross-hybridizing probes were eliminated (30). Upon quality control, 819,799, 818429, and 820,392 probes survived for DNHS, GTP, and NHS, respectively (31). Normalization employed the single-sample Noob (ssNoob) method in minfi (v1.54.1) R package, enabling robust background correction and dye-bias adjustment on a per-sample basis (29). Batch effects due to chip or position were corrected with *ComBat*, protecting key variables age, sex (if applicable), and PTSD status against over-correction (32). The filtered QC’d data was used in further analysis.

### CoRSIV Selection

We focused our analysis on CpG sites located within CoRSIVs (26, 44), genomic regions that exhibit highly consistent DNA methylation patterns across multiple tissues within an individual, spanning ectodermal, mesodermal, and endodermal lineages. CoRSIV genomic coordinates were obtained from published CoRSIVs databases (26, 44). We mapped these regions to the Illumina Infinium MethylationEPIC BeadChip v1 by identifying EPIC array CpG sites whose genomic positions (GRCh37/hg19) fell within CoRSIV boundaries using the manufacturer’s manifest file (v1.0 B5). From this initial set, we retained only CpGs from the EPIC array data in the DNHS that passed quality control, yielding 7,694 CpG sites for subsequent analysis.

### Estimation of covariates

To reduce confounding in DNAm analyses, we adjusted for age, sex, BMI, smoking score, ancestry, and trauma number. Age was categorized into three groups: young (18-34 years), middle-aged (35-54 years), and older (≥55 years) (34). BMI was similarly categorized as normal weight (22-27.9 kg/m²), overweight (28-32.9 kg/m²), and obese (≥33 kg/m²) (35,36). These categorizations were based on previous literature examining age and BMI thresholds in relation to cardiovascular risk profiles (35). DNAm-based smoking scores were computed using established coefficients from 39 CpGs across 27 loci known to capture smoking-associated methylation variation (37). Cellular heterogeneity was addressed by estimating the proportions of major leukocyte subsets (CD4+ T cells, CD8+ T cells, NK cells, B cells, monocytes, and neutrophils) using the Robust Partial Correlation (RPC) method implemented in the EpiDISH R package (38), referencing external datasets (39,40). To account for population stratification, ancestry principal components (PCs) were derived from DNAm data based on CpGs proximal to SNPs identified in the 1000 Genomes Project. Consistent with prior work, PCs 2 and 3, those most associated with self-reported race/ethnicity, were included as covariates (41).

### Statistical Analysis

#### Cardiovascular Disease Prediction Model

To evaluate cardiovascular disease risk predictors and identify DNA methylation biomarkers, we employed a two-tiered analytical approach combining clinical prediction models with epigenome-wide association analyses a methods flowchart provided supplementary Figure 1.

### Clinical Prediction Models

We developed two logistic regression models using 10-fold stratified cross-validation: Model 1 (Base) included age, sex, BMI, trauma number, smoking score, and ancestry principal components; Model 2 added lifetime PTS symptom severity as a continuous variable to test whether psychological trauma burden incrementally predicts CVD risk beyond risk factors and trauma exposure. Age was categorized as 18-34 years, 35-54 years, and ≥55 years (34). BMI categories were normal (22-27.9 kg/m²,), overweight (28-32.9 kg/m²), and obese (≥33 kg/m²) (35,36). We evaluated this hypothesis because prior evidence suggests dose response relationships between PTSS and CVD outcomes, potentially offering better risk stratification than base model (42). Model performance was evaluated using AUC, accuracy, precision, recall, and F1-score. Likelihood ratio tests compared nested models. Stratified 10-fold cross-validation maintained class distribution and prevented overfitting.

### DNA Methylation Association Analyses

To evaluate the independent and adjusted effects of PTSS on CVD risk, we conducted three parallel association analyses using 7,694 CoRSIV-mapped CpG sites: Analysis 1, CVD association without PTSS adjustment and controlling for demographic and lifestyle covariates; Analysis 2, CVD association with PTSS adjustment; and Analysis 3, PTSS severity association using linear regression. For each analysis, separate regression models were fitted for each CpG site. Logistic regression was used for binary CVD outcomes and ordinary least squares for continuous PTSS. Missing values were imputed using mean imputation, and covariates were z-score normalized. Statistical significance was assessed using Benjamini-Hochberg (FDR) correction (threshold: FDR<0.05). As no results survived multiple testing correction, we focused on nominally significant findings (p<0.05) for hypothesis generation. All analyses were conducted using Python and R statistical software (v4.4.2). Additional information is provided in the Supplementary Methods.

### Mediation Analysis

We conducted mediation analysis to assess whether methylation of CpGs implicated in our risk regression models mediates the relationship between PTSS and CVD using the mediation R package (v4.5.1). The analysis estimated the average direct effect (ADE), average causal mediation effect (ACME), and total effect, with all models adjusted for the same covariates a mediation concept provided supplementary Figure 2. Significance was determined using 1000 bootstrap simulations, the default recommended by the package developers to ensure stable uncertainty estimates (43). CpG sites with nominally significant mediation effects were retained for further biological interpretation, providing insight into the potential epigenetic mechanisms linking psychological stress and cardiovascular disease.

### Gene Ontology Enrichment Analysis

We conducted functional enrichment analyses using the missMethyl R package (v1.42.0) (44) on mediator CpG sites, using all 7,694 CoRSIV-mapped CpGs as the background reference set to account for the number of CpG sites per gene on the EPIC array. Gene Ontology (GO) enrichment was assessed across all three ontology categories: Biological Process (BP), Molecular Function (MF), and Cellular Component (CC). KEGG pathway enrichment analysis was performed to identify biochemical pathways associated with the mediator CpGs. All enrichment analyses used the gometh function, which corrects for the varying number of CpG probes per gene. Results were adjusted for multiple testing using the Benjamini-Hochberg false discovery rate (FDR) method with a 5% false discovery rate threshold. This approach enabled the identification of biological processes, molecular functions, cellular locations, and biochemical pathways potentially linked to both trauma exposure and cardiovascular disease through differential methylation.

### Code availability

The scripts generated to perform Cardiovascular Disease Risk Prediction Model, Methylation marker association model associated with CVD, Mediation analysis and Enrichments analysis are available in GitHub (https://github.com/uddin-research-group-at-usf/CorSIVs_Project).

## Results

Table 1 presents the demographic and clinical characteristics of the three cohorts included in this analysis. Our discovery cohort, DNHS (n=523), was predominantly 91% African American participants, a mean age of 54.62 years (SD=16.52), and 60% female. The NHS cohort (n=793) consisted entirely of female nurses (100%), and predominantly of European ancestry (97.2%), with a mean age of 45.05 years (SD=5.57). The GTP cohort (n=601) was primarily African American (94%), with a mean age of 42.59 years (SD=12.02), and 71% female.

**Table 1:**
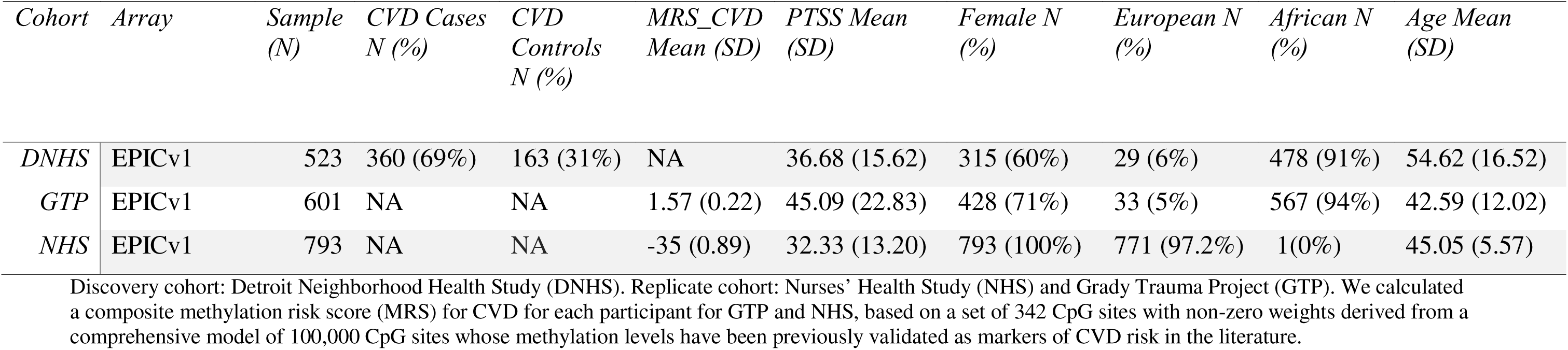
Details overview of the Cohorts.

| *Cohort* | *Array* | *Sample*<br>*(N)* | *CVD Cases*<br>*N (%)* | *CVD*<br>*Controls*<br>*N (%)* | *MRS_CVD*<br>*Mean (SD)* | *PTSS Mean*<br>*(SD)* | *Female N*<br>*(%)* | *European N*<br>*(%)* | *African N*<br>*(%)* | *Age Mean*<br>*(SD)* |
| --- | --- | --- | --- | --- | --- | --- | --- | --- | --- | --- |
| *DNHS* | EPICv1 | 523 | 360 (69%) | 163 (31%) | NA | 36.68 (15.62) | 315 (60%) | 29 (6%) | 478 (91%) | 54.62 (16.52) |
| *GTP* | EPICv1 | 601 | NA | NA | 1.57 (0.22) | 45.09 (22.83) | 428 (71%) | 33 (5%) | 567 (94%) | 42.59 (12.02) |
| *NHS* | EPICv1 | 793 | NA | NA | -35 (0.89) | 32.33 (13.20) | 793 (100%) | 771 (97.2%) | 1(0%) | 45.05 (5.57) |

Discovery cohort: Detroit Neighborhood Health Study (DNHS). Replicate cohort: Nurses' Health Study (NHS) and Grady Trauma Project (GTP). We calculated a composite methylation risk score (MRS) for CVD for each participant for GTP and NHS, based on a set of 342 CpG sites with non-zero weights derived from a comprehensive model of 100,000 CpG sites whose methylation levels have been previously validated as markers of CVD risk in the literature.

### Cardiovascular disease risk assessments

Three multivariable logistic regression models were compared for cardiovascular disease risk prediction (Figure 1). Model 1 (Base Model, standard risk factors only) achieved a cross-validation AUC of 0.664 ± 0.041. Model 2 (Base + PTSS) demonstrated improved performance with an AUC of 0.673 ± 0.043, significantly outperforming the base model (LR = 5.37, p = 0.020). In the best-performing model (Model 2), significant independent predictors of cardiovascular disease included age 55+ years (OR = 2.83, 95% CI: 2.05-3.92, p = 3.40e-10), age 35-54 years (OR = 1.60, 95% CI: 1.18-2.16, p = 0.002), and lifetime PTSD severity (OR = 1.29, 95% CI: 1.03-1.61, p = 0.024) (Table 2). Cross-validation confusion matrices, ROC curves, and comprehensive performance metrics confirmed Model 2’s superior predictive accuracy across all evaluated metrics (Figure 1). These findings demonstrate that continuous PTSS provides better predictive value for cardiovascular disease risk than PTSD diagnosis, even after controlling for trauma exposure and standard cardiovascular risk factors.

**Figure 1:**
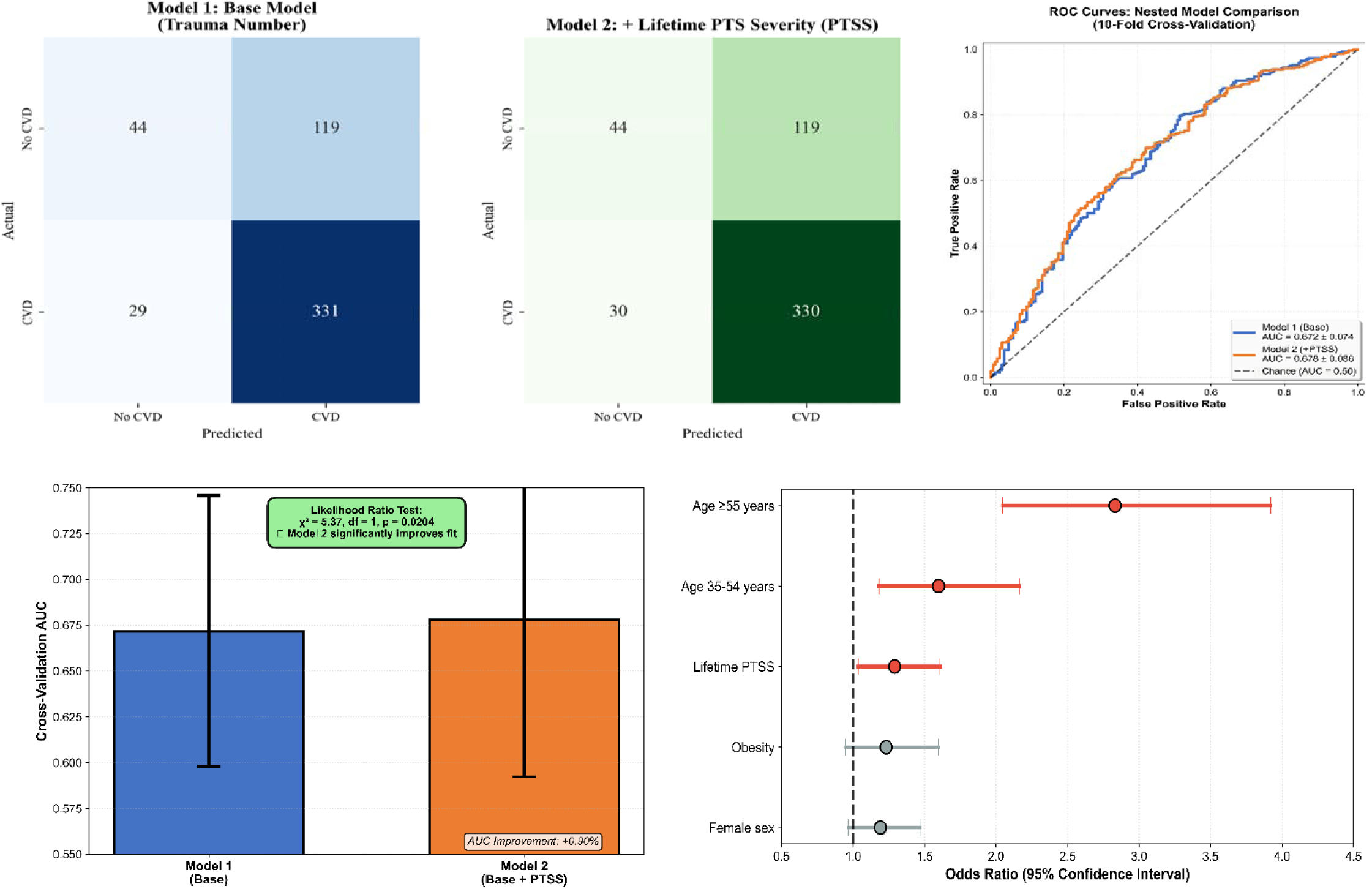
Predictive Performance of Trauma and PTSD-Based Models for Cardiovascular Disease Risk in the DNHS Cohort (N=523). Two logistic regression models were evaluated using 10-fold cross-validation: Model 1 (trauma number), and Model 2 (lifetime PTS symptom severity), Confusion matrices (top left 2 panels), ROC curves (top right). The likelihood ratio test (bottom left) indicates that Model 2 significantly outperforms the base model (p = 0.028). Forest plot (bottom right) displays odds ratios for key CVD risk factors in Model 2. All models adjusted for age, sex, BMI, and ancestry component.

**Table 2:** Cardiovascular disease risk assessment across model.

| *Risk Factor* | *Model 1* |  |  |  |  |  | *Model 2* |  |  |  |  |  |
| --- | --- | --- | --- | --- | --- | --- | --- | --- | --- | --- | --- | --- |
|  | *Coefficient* | *Std Err* | *Z* | *OR* | *95% CI* | *p-value* | *Coefficient* | *Std Err* | *Z* | *OR* | *95% CI* | *p-value* |
| *Age 35-54 years* | 0.473 | 0.1537 | 3.0779 | 1.6 | 1.19-2.17 | 0.002* | 0.468 | 0.1545 | 3.0296 | 1.6 | 1.18-2.16 | 0.002* |
| *Age ≥ 55 years* | 1.0236 | 0.1644 | 6.2271 | 2.78 | 2.02-3.84 | 4.75E-10* | 1.0408 | 0.1658 | 6.2792 | 2.83 | 2.05-3.92 | 3.40E-10* |
| *Overweight (BMI 28-32.9)* | 0.1265 | 0.1323 | 0.9559 | 1.13 | 0.88-1.47 | 0.339 | 0.1247 | 0.1334 | 0.9345 | 1.13 | 0.87-1.47 | 0.35 |
| *Obese (BMI ≥33)* | 0.2168 | 0.1323 | 1.6379 | 1.24 | 0.96-1.61 | 0.101 | 0.2077 | 0.1329 | 1.5625 | 1.23 | 0.95-1.60 | 0.118 |
| *Number of traumas* | 0.1561 | 0.1098 | 1.4221 | 1.17 | 0.94-1.45 | 0.155 | 0.0989 | 0.1131 | 0.8739 | 1.1 | 0.88-1.38 | 0.382 |
| *Female (sex)* | 0.2016 | 0.105 | 1.9197 | 1.22 | 1.00-1.50 | 0.055 | 0.175 | 0.1063 | 1.6466 | 1.19 | 0.97-1.47 | 0.1 |
| *smoking score* | 0.1547 | 0.1054 | 1.467 | 1.17 | 0.95-1.44 | 0.142 | 0.1494 | 0.106 | 1.4095 | 1.16 | 0.94-1.43 | 0.159 |
| *Ancestry PC2* | 0.1799 | 0.0964 | 1.866 | 1.2 | 0.99-1.45 | 0.062 | 0.1625 | 0.0972 | 1.6714 | 1.18 | 0.97-1.42 | 0.095 |
| *Ancestry PC3* | -0.116 | 0.1023 | -1.1345 | 0.89 | 0.73-1.09 | 0.257 | -0.1217 | 0.1028 | -1.1835 | 0.89 | 0.72-1.08 | 0.237 |
| *Lifetime PTSS* | — | — | — | — | — | — | 0.2548 | 0.1128 | 2.2589 | 1.29 | 1.03-1.61 | 0.024* |
|  | *Model Performance* |  |  |  |  |  | *Model Performance* |  |  |  |  |  |
| *Cross-validation AUC (mean ± SD)* | 0.680 ± 0.074 |  |  |  |  |  | 0.683 ± 0.086 |  |  |  |  |  |
| *AIC* | 606.5 |  |  |  |  |  | 603.1 |  |  |  |  |  |
| *Likelihood ratio test vs Model 1* | — |  |  |  |  |  | P= 0.020* |  |  |  |  |  |

The CVD risk factor assessments were tested across all possible main covariates. Age (Mid and old age), Sex (Female), BMI (Overweight and Obese), PTSS (Life PTS severity), Traum exposure (Traumanum), Smoking score (SmoS), and Ancestry principal components (Comp.2 and Comp.3). For each risk factor prediction model was done with regression analysis with nominal significant score (\* p < 0.05), Odd ratio, and 95% cofinance interval. AUC = Area Under the Curve; AIC = Akaike Information Criterion; BIC = Bayesian Information Criterion, LR test = Likelihood Ratio test comparing each model to Model 1 (Base), Reference categories: Age 18-34, Normal BMI, Male sex, all continuous variables are standardized. Cross-validation performed with 10-fold stratified sampling with nominal significant score (\* p < 0.05).

**Table 3.** CpG sites that mediate the relationship between PTSD symptom severity (PTSS) and cardiovascular disease (CVD) across cohorts.

| *CpGs* | *Gene* | *Chr* | *DNHS Cohort* |  |  | *GTP Cohort* |  |  | *NHS Cohort* |  |  | *Status* |
| --- | --- | --- | --- | --- | --- | --- | --- | --- | --- | --- | --- | --- |
|  |  |  | ACM<br>EP-value | Prop.<br>Mediat<br>ed | ACME<br>Directio<br>n | ACME<br>P-value | Prop.<br>Mediated | ACME<br>Direction | ACME<br>P-value | Prop.<br>Mediated | ACME<br>Direction |  |
| *cg00817557* | *MIR646*<br>*HG* | 20 | *0.006 | -0.194 | -0.00059 | 0.27 | -0.294 | -0.00074 | 0.59 | 0.124 | -0.0013 | Shared<br>CpGs<br>across<br>all m<br>odels |
| *cg01160818* | *C5orf56* | 5 | *0.04 | -0.12 | -0.0004 | 0.76 | -0.057 | -<br>0.000145 | 0.51 | -0.09 | -0.0017 |  |
| *cg02841227* | *HIST1H*<br>*4A* | 6 | *0.038 | 0.102 | 0.0032 | 0.33 | -0.216 | -0.00054 | 0.53 | -0.053 | -0.0016 |  |
| *cg07941916* | *C5orf56* | 5 | *0.016 | 0.137 | 0.00044 | 0.39 | 0.66 | 0.001654 | 0.51 | -0.099 | -0.0017 |  |
| *cg17459387* | *AATK* | 17 | *0.032 | -0.134 | -0.00041 | 0.85 | -0.024 | -<br>0.000061 | 0.50 | -0.106 | -0.0017 |  |
| *cg20545458* | - | 6 | *0.042 | 0.127 | 0.0004 | 0.73 | 0.202 | 0.000504 | 0.43 | -0.17 | -0.0018 |  |
| *cg25181130* | *RHBDL*<br>*3* | 17 | *0.048 | -0.134 | -0.0004 | 0.56 | 0.64 | 0.001603 | 0.41 | -0.19 | -0.0018 |  |

The table presents CpG sites identified as nominally significant mediators of the PTSS–CVD association in the discovery cohort, and their corresponding results in each additional replication cohort. For each site, the average causal mediation effect (ACME), proportion mediated, and direction of effect are reported. Asterisks (\*) indicate nominal significance at $p < 0.05$ .

### Association between DNA methylation and cardiovascular disease risk with and without PTSS

Our analysis identified 413 nominally significant CpG sites associated with CVD when accounting for PTSS and trauma exposure (Figure 2; Analysis 1). When PTSS was excluded from the association analysis (Figure 2; Analysis 2), 405 CpG sites showed nominally significant associations with CVD. Additionally, 490 CpG sites were associated with PTSS (Figure 2; Analysis 3). Comparative analyses revealed 22 overlapping CpG sites between Analysis 1 and 3, as well as 22 overlapping sites between Analysis 2 and 3. Across the three analyses, we identified 27 unique CpGs: 17 appeared in the overlaps among the three models, while 5 were unique to Analysis 1 and 5 were unique to Analysis 2. Notably, no associations survived FDR correction in the discovery cohort.

**Figure 2.**
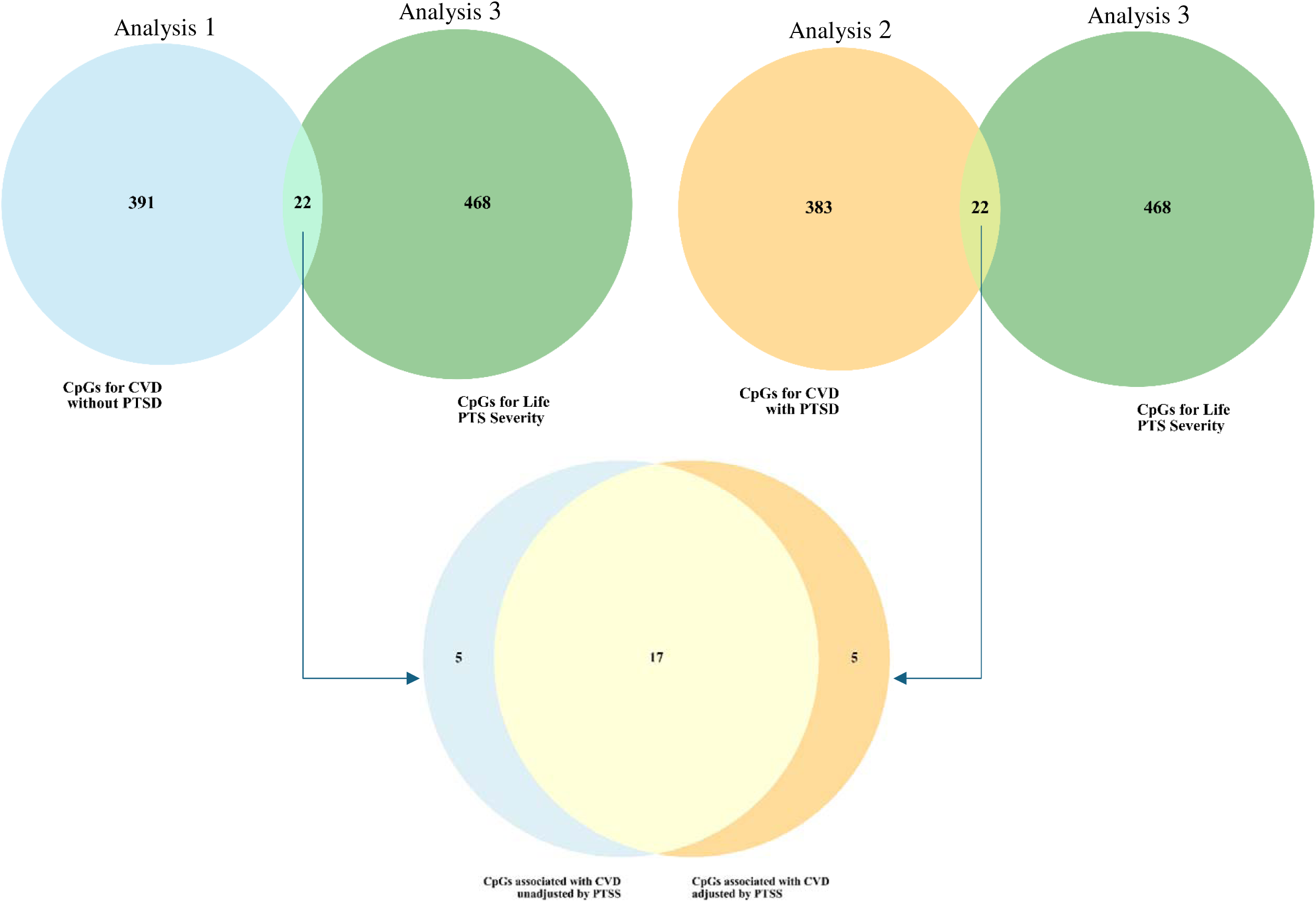
Overlap of methylation sites associated with PTSS and CVD across regression models in the DNHS Cohort (N=523). The Venn diagram illustrates the overlap of CpG sites identified across three models: Analysis 1: CpGs associated with CVD in models unadjusted (top left sky blue) and Analysis 2: adjusted (top right orange) for PTSD symptom severity (PTSS), and Analysis 3: CpGs associated with PTSS alone (green). The bottom portion of the figure displays the subset of CpGs unique to or shared between each comparison group.

The majority (approximately 60%) of the 27 CpG sites potentially associated with both PTSS and CVD risk exhibited positive associations, suggesting that higher methylation at these loci may be linked to increased cardiovascular risk in trauma-exposed individuals. The remaining sites (40%) showed negative associations (Supplementary Figure 3).Additionally, Regression analyses confirmed that methylation levels at these CpG sites correlated with age, consistent with known age-related methylation drift. To explore potential biological mechanisms linking trauma exposure to cardiovascular outcomes, we next examined whether these 27 identified CpG sites might statistically mediate the relationship between PTSS and CVD risk.

Mediation analysis revealed a subset of these 27 CpG sites that potentially mediate the relationship between PTSS and CVD risk. Table 4 presents mediating CpGs with nominally significant effects in the discovery cohort, along with the corresponding results in the two replication cohorts. In the discovery cohort, seven of the 27 CpG sites showed nominally significant (p < 0.05) average causal mediation effects (ACME) between PTSS and CVD (Table 4). Three of the mediating CpGs in the discovery cohort exhibited positive ACME values, indicating that increased PTSD symptom severity was associated with methylation differences that, in turn, corresponded to increased CVD risk (45). Examining directional consistency across cohorts, six of seven mediating CpGs showed effects in the same direction in at least one replication cohort. In GTP, five of the seven CpGs (cg00817557, cg01160818, cg07941916, cg17459387, and cg20545458) had mediation effects in the same direction as in DNHS, whereas in NHS four CpGs were directionally consistent, all in the negative direction (Table 4). Three CpGs showed concordant mediation directions across all three cohorts. Notably, cg07941916 (C5orf56) and cg20545458 demonstrated positive ACME values in both DNHS and GTP (Figure 3), suggesting directionally consistent, risk-enhancing effects despite not reaching nominal significance in the replication cohort (p = 0.39). In the NHS, the uniformly negative ACME directions indicate a different pattern of mediation than in DNHS, which likely reflects the NHS cohort’s stringent health-based selection criteria (including no smoking in the past 10 years) and its distinct demographic profile compared with the predominantly urban DNHS and GTP populations. Complete mediation results for all analyzed CpG sites, including ACME, Average Direct Effects (ADE), and total effects with their respective 95% confidence intervals across all three cohorts, are provided in Supplementary Table S1.

**Figure 3.**
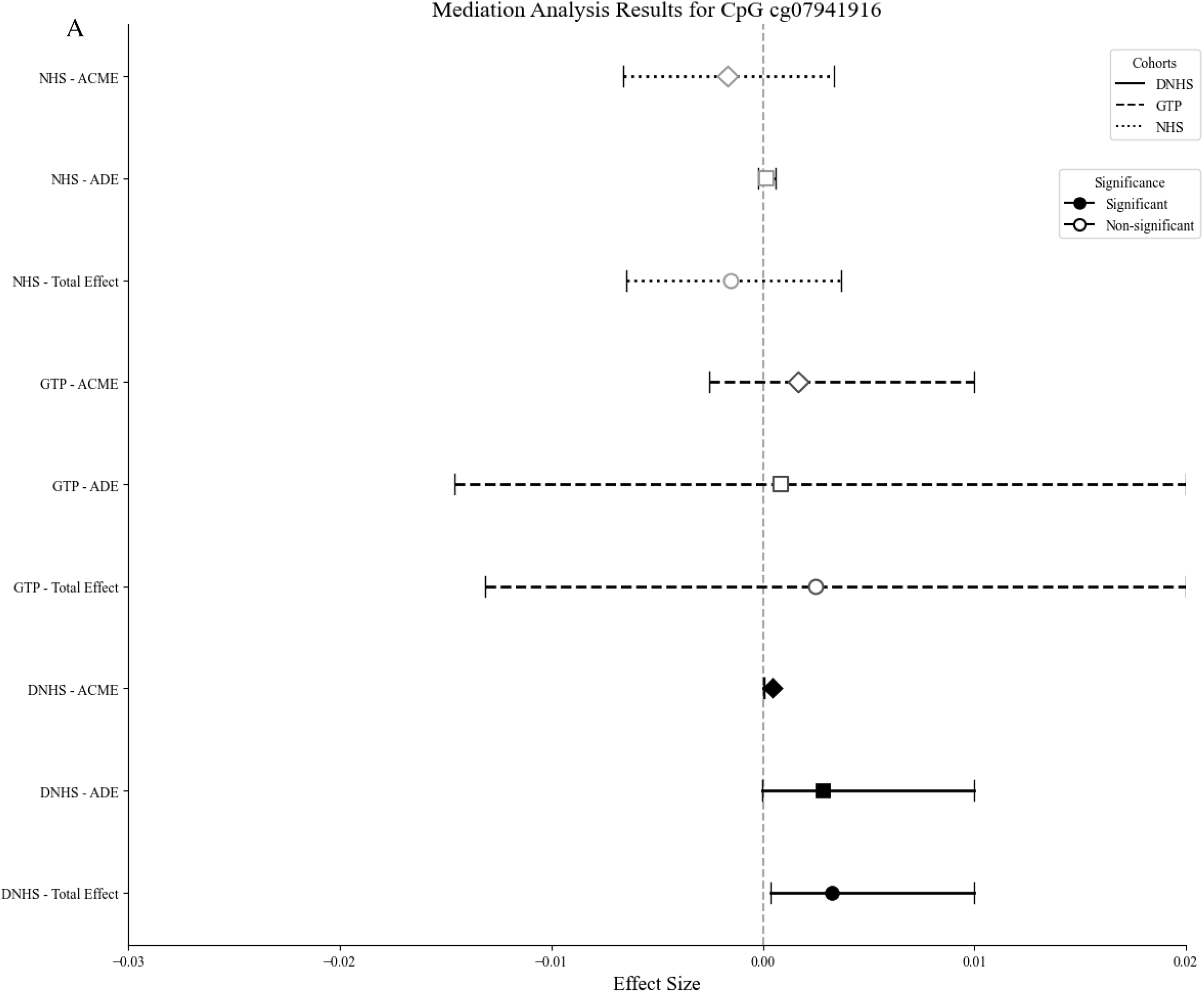
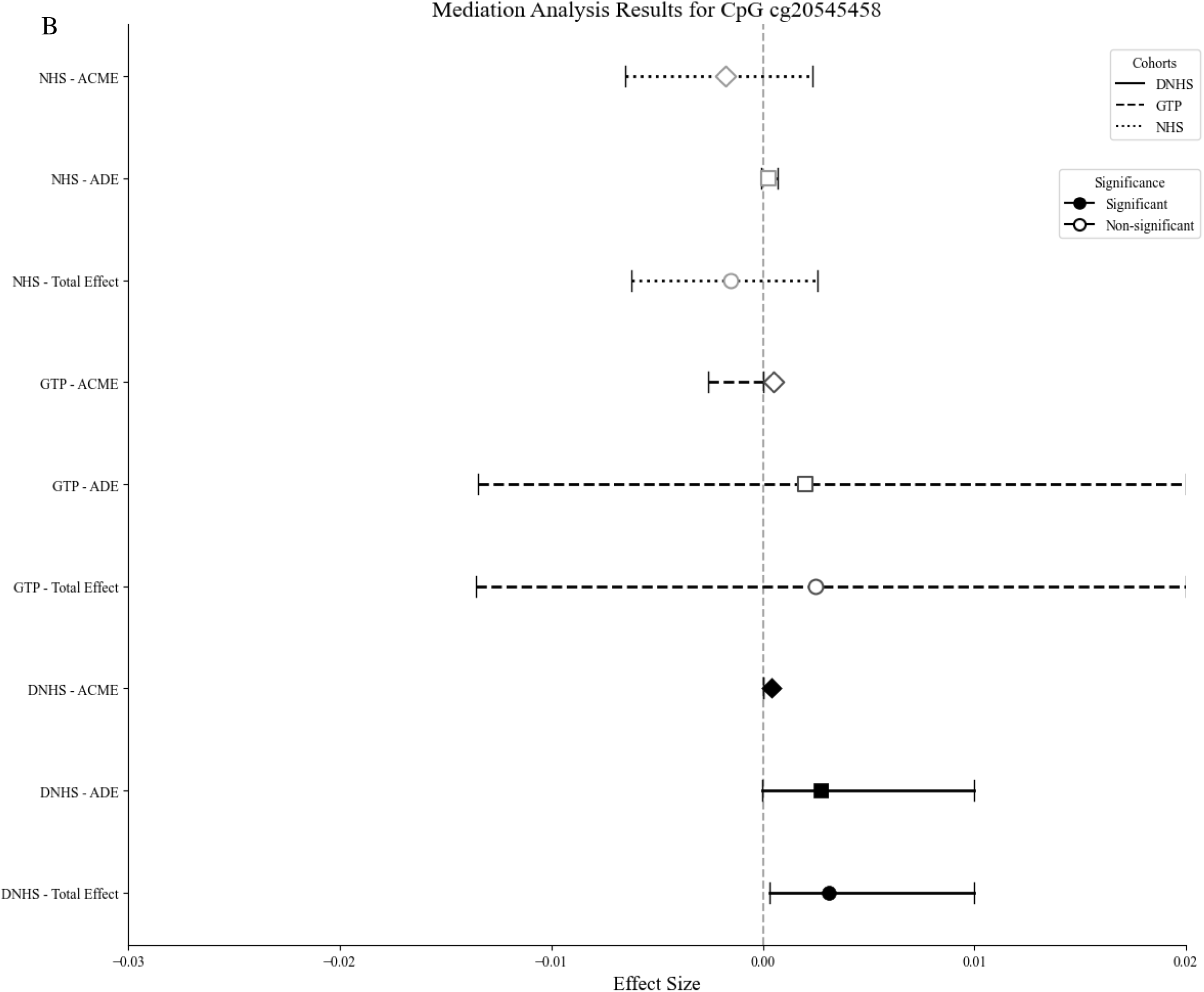
Forest plot of nominally significant mediation effects for a cross-cohort CpG mediator on the DNHS Cohort (N=523), GTP (n=601), and NHS (n = 793). This figure displays A. the mediation effect of CpG site cg07941916, and B. the mediation effect of CpG site cg20545458 found to be a nominally significant mediator (*p* < 0.05) of the PTSS–CVD association in the discovery and a replication cohort with direction consistency. The forest plot shows the average causal mediation effect (ACME), Indirect Effect-ACME in a diamond shape, the average direct effect (ADE) in a rectangular shape, and the Total Effect in a circle shape, along with 95% confidence intervals for each cohort. Filled shapes represent significant mediation effects; open shape indicates non-significant effects.

### Functional Pathway Analysis of Identified CpG Sites

Gene Ontology over-representation (missMethyl) of the seven mediating CpGs showed nominal enrichment (p<0.05) for terms related to telomere/chromatin biology (telomere organization, nucleosome assembly, structural constituent of chromatin), protease activity (serine/serine-type peptidase/hydrolase activity), protein localization to chromosome, and megakaryocyte differentiation (Figure 4 and Supplementary Table S3). No terms survived multiple-testing correction (FDR). Given the limited number of CpGs analyzed (n=7), this enrichment results should be interpreted cautiously as hypothesis-generating observations that require validation with larger CpG sets in independent cohorts.

**Figure 4:**
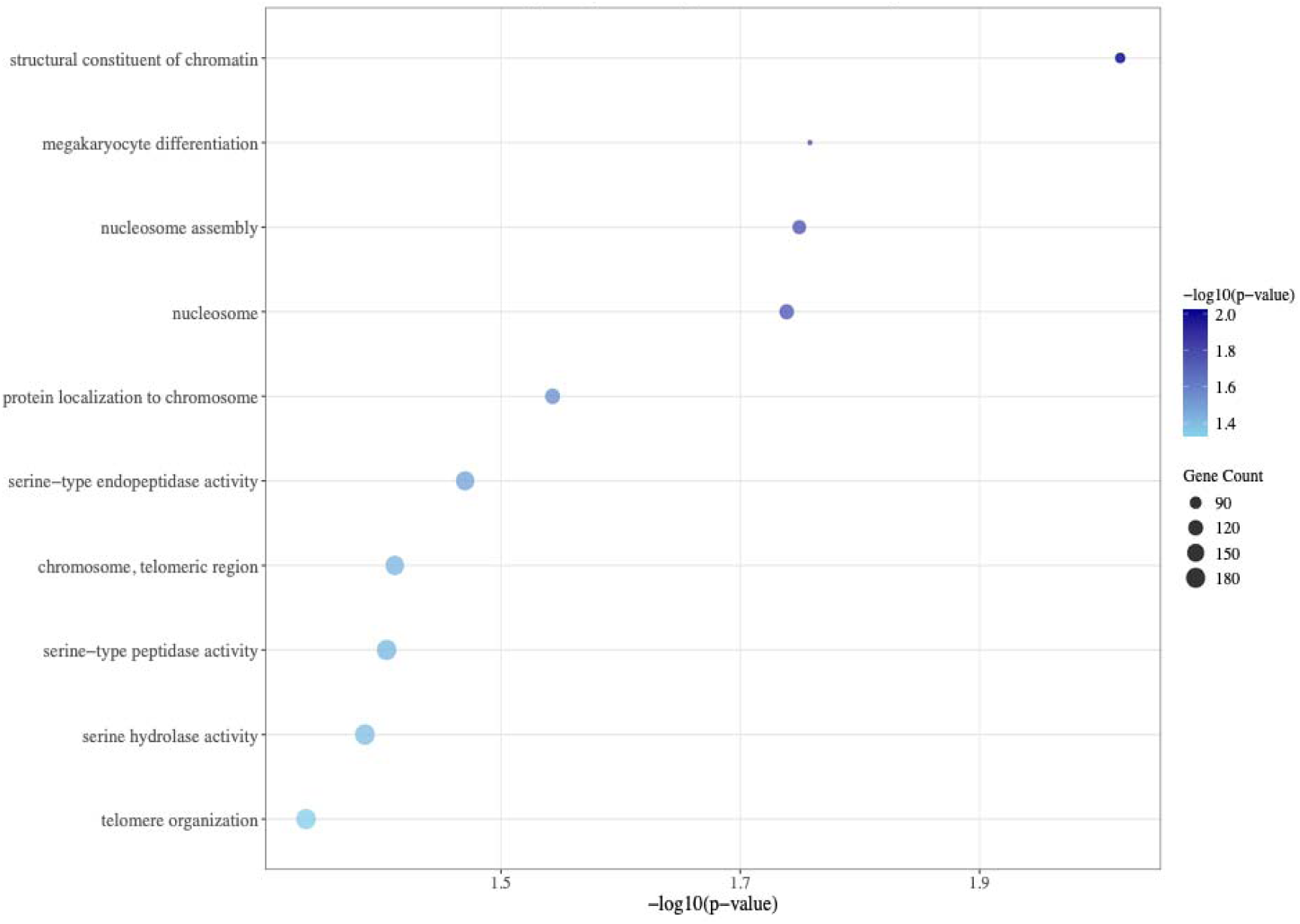
Pathway enrichments of the CpG sites associated with CVD in the DNHS Cohort (N=523). Gene Ontology pathway enrichment analysis of seven CVD-associated CpG sites from the Discovery cohort. Analysis used missMethyl R package with CoRSIVs (7694 CpGs) as background. The dot plot shows top 10 enriched GO terms (y-axis) versus significance (-log10(p-value), x-axis). Dot color intensity indicates significance level (darker blue = higher significance), while size represents gene count (90-180 genes). Pathways related to chromatin structure (most nominally significant, p=0.01, 150 genes), nucleosome assembly, and serine-type peptidase activity were nominally enriched, suggesting epigenetic mechanisms linking PTSS to cardiovascular risk.

## Discussion

In our discovery cohort (DNHS), we identified seven CpG sites within conserved genomic regions (CoRSIVs) where methylation levels nominally mediated the association between post traumatic stress symptoms and cardiovascular risk. Across these seven CpGs, mediation effects did not reach statistical significance in the replication cohort, but we observed a notable pattern in effect directions: in GTP, five of the seven CpGs showed effects in the same direction as in DNHS, and in NHS four CpGs were directionally consistent, yielding three CpGs with concordant mediation directions across all three cohorts. This pattern suggests some stability of the methylation-CVD relationship across cohorts, while also indicating context dependence. Within this directionally consistent set, cg07941916 (C5orf56/IRF1 AS1) and cg20545458 (intergenic) showed positive mediation in DNHS with the same direction in GTP and supportive exploratory functional and genetic annotations. Exploratory cis meQTL analyses in external cohorts indicated that these two CpGs (cg07941916 and cg20545458) are influenced by nearby genetic variants with large effects, with cg07941916 additionally showing cross ancestry, directionally consistent cis effects in GENOA and EPIGEN (Details in Supplementary File and Supplementary Table 2). GWAS look-ups for the lead cis-SNPs in the Freeze 3 datasets (46), however, revealed only nominal or absent associations with PTSD; therefore, the genetic evidence for these loci is best viewed as supportive but modest. Taken together, the combination of three cohort directional consistency and exploratory genetic regulation suggests that cg07941916 and cg20545458 warrant further investigation, while the remaining CpGs should be treated as exploratory.

The differential replication patterns across our cohorts reflect their distinct demographic profiles. Both DNHS (91% African American) and GTP (94% African American) represent urban populations with high trauma exposure and smoking history (31,47), whereas NHS participants (97% European ancestry) comprise a relatively healthy, well-educated group with lower baseline CVD prevalence (48). The NHS cohort was included to evaluate whether associations identified in high-risk, trauma-exposed populations extend to lower-risk groups with different ancestry and environmental backgrounds. Therefore, the attenuated or reversed effects observed for loci such as cg07941916 (C5orf56) and cg20545458 underscore the importance of considering population context when interpreting trauma-related epigenetic associations.

Our findings both complement and extend previous research on the biological embedding of psychological trauma. The connection between psychological trauma and heart disease has been documented by Pollard et al. (2016), who observed significant links between combat trauma and cardiovascular illness (49). Blood-based DNAm studies indicate that traumatic stress and PTSD are associated with immune-related DNAm differences, with injury cohorts showing pathway enrichment in immune signaling and PTSD meta-analyses identifying immune-linked loci (28,50). In complementary preclinical work, Zhao et al. (2020) showed that stress can repress brain-derived neurotrophic factor pathways via H3K9me2 in rat hippocampus (51), one plausible epigenetic route by which stress affects the brain (52).

In the discovery cohort, seven CpGs showed nominal mediation effects, including some with plausible links to immune, stress-response, or cardiovascular pathways. Notably, cg07941916 is annotated to C5orf56 (IRF1-AS1/CARINH), a long non-coding RNA involved in innate-immune regulation (IRF1; interferon/NF-κB) (53). Dysregulation of innate immune signaling has been previously reported in PTSD (54), including in methylation-based studies (54,55). The C5orf56/IRF1-AS1/CARINH locus acts as a regulatory element modulating expression of the nearby transcription factor called interferon regulatory factor 1 (IRF1), which is critical for hematopoietic stem cell (HSC) maintenance by regulating self-renewal, stress responses, apoptosis, and inflammatory signaling; IRF1 dysregulation is linked to acute myeloid leukemia (AML) (56). This finding aligns with a broader body of evidence linking PTSD to immune dysregulation and cardiovascular risk (15,57,58). PTSD is associated with elevated inflammatory cytokines (e.g., IL-1, IL-6, TNF-α), which can cross the blood–brain barrier and activate microglia, contributing to neuroinflammation (56). Chronic inflammation in PTSD is also linked to increased risk of autoimmune disorders and cardiovascular disease (59). However, alternative and complementary pathways warrant consideration. PTSD is associated with increased use of both prescription medications (e.g., antidepressants, anxiolytics, antipsychotics) and illicit substances, many of which can independently alter cardiovascular health and potentially influence DNA methylation patterns. Thus, the observed methylation changes could reflect medication or substance use effects rather than, or in addition to, direct biological consequences of PTSD (21). Further supporting a role for inflammation-related epigenetic markers, the other directionally consistent CpG, cg20545458, has been robustly associated with chronic obstructive pulmonary disease (COPD), another condition characterized by chronic inflammation(60).

Another notable mediator, cg17459387, is located within the Apoptosis-associated tyrosine kinase (*AATK)* gene region. While *AATK* has been studied primarily in cancer, neuronal contexts, and Alzheimer’s dementia (61,62), it has potential localization to mitochondria and is involved in brain development (61,63). AATK is predominantly known for its roles in neuronal development and apoptosis, including implications in neurodegenerative diseases (64).

Likewise, we observed a few mediators align with stress-response and aging pathways. A robust body of evidence links PTSD to an increased risk of CVD through complex biological pathways involving inflammation, autoimmune, and aging. For instance, cg25181130 is located in the *RHBDL3* gene (a member of the rhomboid protease family) and has been reported as a highly age-associated CpG site in blood (65). This suggests that methylation at *RHBDL3* changes with chronological age and may capture aspects of biological aging or cumulative stress exposure. PTSD has been linked to accelerated epigenetic aging and premature onset of age-related conditions, but its age-linked methylation suggests a broader stress/aging signal in trauma-exposed populations (66,67). At the gene-family level, rhomboid proteases can affect cardiac function in model organisms (e.g., Drosophila) (68). In our study, both CpGs nominally mediated the association between PTSS and CVD (ACME, p<0.05), making it intriguing that a known age-related epigenetic locus emerged as a PTSS-CVD mediator in our data. A known age-related epigenetic locus emerged as a PTSS-CVD mediator in our data. The CpG cg02841227 annotates to *HIST1H4A* (H4C1); although we did not find direct links between this gene and autonomic control or CVD, histone H3/H4 modifications more generally are implicated in cardiac hypertrophy and heart failure, so we view this site as a chromatin marker rather than a gene-specific cardiac effector (69,70). Finally, with respect to cg00817557, located in the *MIR646HG* long non-coding RNA host gene, we found no direct cardiovascular or autonomic literature for the lncRNA host gene and therefore treat it as exploratory. Overall, this systems view accords with neurovisceral-integration work linking chronic stress, reduced heart-rate variability, and elevated cardiovascular risk (71,72).

Our study has limitations. Its cross-sectional design limits causal interpretation, and although major covariates were adjusted for, residual confounding is possible. Additionally, different MRS_CVD baseline distributions across ethnically and behaviorally distinct populations may influence mediation directions and limit direct cross-cohort comparisons. Peripheral blood DNA may not fully represent tissue-specific methylation patterns relevant to cardiovascular biology; however, focusing on CoRSIV regions enhances biological relevance across tissues. While our focus on CoRSIVs enhances the cross-tissue relevance of our findings, future studies should incorporate tissue-specific validation in cardiovascular tissues where ethically feasible. Additionally, longitudinal designs would help establish temporal relationships between trauma exposure, epigenetic changes, and cardiovascular outcomes. Replication across independent and diverse cohorts remains a challenge, underscoring the need for larger harmonized studies. Future research should incorporate longitudinal designs and functional assays, including multi-omics integration, to clarify causality and identify actionable therapeutic targets.

### Conclusion

Our findings provide preliminary evidence that DNAm within CoRSIV regions may partially mediate the association between PTSS and CVD risk, based on seven nominal mediators in DNHS and three CpGs showing concordant mediation directions across all three cohorts, with six of the seven displaying directionally consistent effects in at least one replication cohort. Within this directionally consistent set, cg07941916 and cg20545458 showed positive mediation in DNHS with the same direction in GTP; and supportive, but exploratory, functional and genetic annotation analyses indicate that these CpGs reside in genetically regulated loci with links to inflammatory and cardiometabolic traits. In contrast, the uniformly negative mediation effects observed in NHS likely reflect its healthier, lower-risk demographic profile, underscoring population-specific trauma-related epigenetic signatures rather than definitive non-replication. Overall, these signals, spanning immune, neuronal/stress-aging, and autonomic pathways, are hypothesis-generating and require validation in larger, ancestrally diverse longitudinal cohorts with functional follow-up before any clinical interpretation.

## Declaration of generative AI and AI-assisted technologies in the manuscript preparation process

During the preparation of this work, the author(s) used generative AI-assisted technologies (e.g., Grammarly, Nature AI Research Assistant, and Perplexity) in order to improve grammar, clarity, and sentence structure. After using these tools, the author(s) reviewed and edited the content as needed and take full responsibility for the scientific integrity and content of the published article.

## Data Availability

The DNHS data is available at dbGAP (phs000560.v2.p1). The GTP data is available at GSE132203 in the GEO database. The NHS data is available via an approved data use request to NHS investigators.

## Acknowledgement

We gratefully acknowledge the support from the National Institute of Health (NIH; R01MD011728; R01MH108826). We also thank the team of Nurses’ Health Study (NHS) for their valuable support and technical assistance. More grants to be added before sharing with co-authors.

