## Supplementary Methods for "DNA Methylation as a Mediator of Cardiovascular Disease Risk in Relation to PTSD Severity: Identification of Potential Epigenetic Biomarkers"

PTSD - Posttraumatic Stress Disorder, Cardiovascular Disease, Biomarkers, Physical Health

**Corresponding author:**

Address correspondence to:

Monica Uddin, PhD

University of South Florida

3720 Spectrum Blvd., Suite 304

813-974-9765

### Statistical Analysis Methods Flowchart

The research design with multiple steps includes trait risk assessments to methylation weight-based disease prediction, as well as sample preparation, including sample selection, estimation of covariates Supplementary Figure 1.

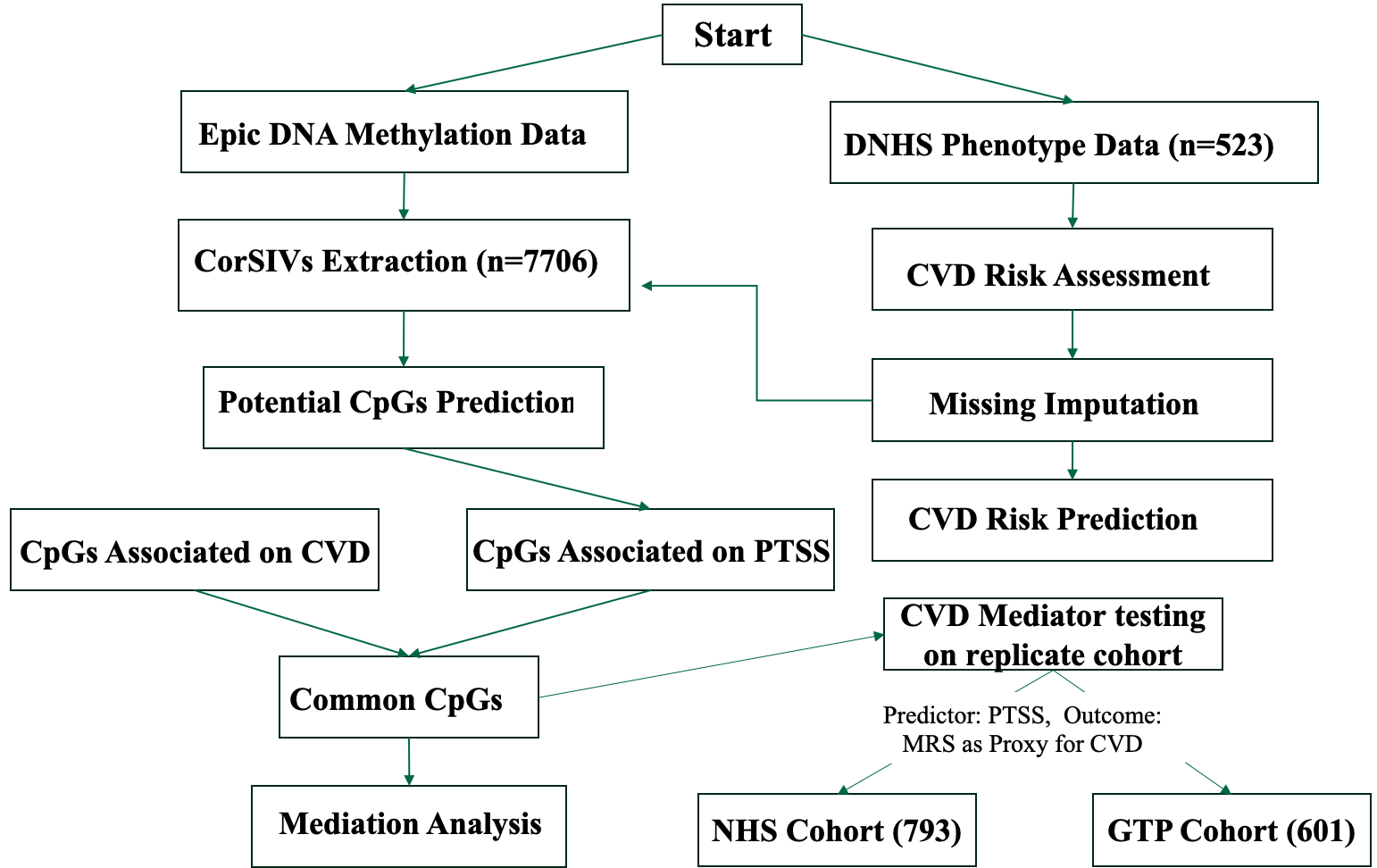

**Supplementary Figure 1: Research Design and Methods flowchart.**

### Calculation of Methylation Risk Score (MRS_CVD):

We analyzed data from three cohorts: the Detroit Neighborhood Health Study (DNHS), Nurses' Health Study (NHS), and Grady Trauma Project (GTP). For the replication cohorts (GTP and NHS), which lacked direct cardiovascular disease (CVD) measures, we computed a methylation risk score (MRS_CVD) using the methodology established by Westerman et al. (2020).

We computed a methylation risk score (MRS) as an indicator of cardiovascular disease (CVD) risk utilizing DNA methylation data. We employed the proven methodology established by Westerman et al., 2020, utilizing coefficients accessible on GitHub (<https://github.com/kwesterman/meth_cvd/blob/master/output/mrs_calculation/combined_mrs_coefs.rds>).

The MRS for each participant was computed using the formula

$$MRS_{CVD}= \sum_{i=1}^{n} (\boldsymbol{\beta i}\times\boldsymbol{wi})$$

β_i_ = Methylation levels for specific CpG sites.

*W_i_* = Predefined weights from Westerman et al.

*n* = Total number of CpG sites included.

### Risk Prediction in Discovery Cohort (DNHS):

A multivariable logistic regression approach was implemented in the discovery cohort (DNHS) to examine the association between post-traumatic stress symptom (PTSS) severity and cardiovascular disease (CVD) status. We developed three nested models with progressively added complexity to assess the independent contribution of PTSD-related variables while controlling for established cardiovascular risk factors (1).

**Outcome Definition**

CVD was defined as the presence of self-reported or physician-diagnosed hypertension or heart-related conditions, including chest pain, history of rheumatic fever, or irregular heart rhythms (arrhythmias). This definition is consistent with clinical criteria for identifying individuals at elevated cardiovascular risk and has been validated in similar epidemiological studies (1). Participants were classified as CVD cases if they reported any of these conditions at the time of assessment.

**Predictor Variables**

The primary predictor of interest was lifetime PTSS severity, assessed using a lifetime-adapted version of the PTSD Checklist-Civilian version (PCL-C). Unlike the standard PCL-C which assesses symptoms over the past month, this modified version measured lifetime symptom severity. PTSS was treated as a continuous variable in Model 2 and as a binary diagnosis (PTSD present/absent) in Model 3.

**Covariates**

All models included the following covariates:

Age: Categorized into three groups based on cardiovascular risk profiles: 18-34 years (reference, low risk), 35-54 years (intermediate risk), and ≥55 years (elevated risk) (2). Sex: Biological sex (male/female) was included, given well-established sex differences in cardiovascular disease prevalence and presentation. Body Mass Index (BMI): Calculated as weight (kg)/height (m²) and categorized as normal (22-27.9 kg/m², reference), overweight (28-32.9 kg/m²), and obese (≥33 kg/m²), following established cutoffs for African American populations (3,4). Smoking Score: The study used DNAm-based smoking scores were computed using established coefficients from 39 CpGs across 27 loci known to capture smoking-associated methylation variation (5).Ancestry Principal Components: To account for population stratification, ancestry principal components (PCs) were derived from DNAm data based on CpGs proximal to SNPs identified in the 1000 Genomes Project. Consistent with prior work, PCs 2 and 3, those most associated with self-reported race/ethnicity, were included as covariates (6)

**Model Specifications**

Model 1 (Base Model): CVD ~ Age Categories + Sex + BMI Categories + Smoking Score + Trauma Number + Ancestry PCs (PC2 + PC3)

This base model evaluated standard cardiovascular risk factors without PTSD-related variables to establish baseline predictive performance.

Model 2 (PTSS Severity Model): CVD ~ Age Categories + Sex + BMI Categories + Smoking Score + Trauma Number + Ancestry PCs (PC2 + PC3) + Lifetime PTSS (Continuous)

Model 2 added lifetime PTSS severity as a continuous predictor to assess whether psychological trauma burden incrementally predicts CVD risk beyond traditional risk factors.

Model 3 (Binary PTSD Model): CVD ~ Age Categories + Sex + BMI Categories + Smoking Score + Trauma Number + Ancestry PCs (PC2 + PC3) + Lifetime PTSD (Binary)

Model 3 substituted binary PTSD diagnosis for continuous severity to determine whether clinical diagnostic threshold provides equivalent or superior predictive value compared to dimensional severity.

**Model Validation and Performance Assessment**

All models were validated using stratified 10-fold cross-validation to ensure robust performance estimates and prevent overfitting. Stratification maintained the proportion of CVD cases and controls within each fold, addressing class imbalance. For each fold, 90% of the data was used for training and 10% for testing, with this process repeated 10 times to ensure all observations served as test data exactly once.

**Model Comparison**

Nested models were compared using likelihood ratio tests to determine whether the addition of PTSD-related variables significantly improved model fit. The test statistic follows a chi-square distribution with degrees of freedom equal to the difference in the number of parameters between models. Statistical significance was assessed at α = 0.05.

**Statistical Software**

All analyses were conducted using R statistical software (version 4.4.2) with the following packages: stats (base logistic regression), caret (cross-validation), pROC (ROC curve analysis), lme4, and ggplot2 (visualization) (7). Python (version 3.12.1) with statsmodels and scikit-learn libraries (8) was used for supplementary analyses and validation.

**Missing Data**

Missing covariate data were minimal (<5% for all variables). Complete case analysis was used as the primary approach. Sensitivity analyses using multiple imputation confirmed the robustness of findings.

**DNAm-based prediction models**

DNAm-based prediction models in our discovery cohort using 7,694 CpG sites within CoRSIVs. To investigate the role of DNAm, we then ran three separate models. The first model assessed whether CpG methylation levels predicted CVD risk while controlling for standard covariates (age, sex, BMI, smoking status, and trauma exposure) but excluding PTSS. The second model added PTSS as a covariate to determine if including traumatic stress altered these methylation-CVD associations. The third model examined whether methylation patterns predicted PTSS while controlling for the same covariates. In the disease risk prediction model, CpGs were used as primary predictors of CVD, and the focus was on their direct association with disease risk. Since the model aimed to assess predictive power rather than causal pathways, adjusting for cell type proportions was not essential. We used logistic regression for the binary CVD outcome (Models 1 and 2) and linear regression for the continuous PTSS outcome (Model 3). Below is the list of each model.

Model1: CVD ~ CpGs + Age Categories + Sex + BMI Categories + Smoking Score + Trauma number + Ancestry PCs (PC2 + PC3)

Model2: CVD ~ CpGs + PTSS + Age Categories + Sex + BMI Categories + Trauma number + Smoking Score + Ancestry PCs (PC2 + PC3)

Model3 PTSS ~ CpGs + Age Categories + Sex + BMI Categories + Smoking Score + Trauma number + Ancestry PCs (PC2 + PC3)

### Mediation Analysis

The mediation analysis was performed using the mediation (4.5.1) R package, estimating the average direct effect (ADE), average causal mediation effect (ACME), and total effect for each CpG site (9).

**Mediation Model Specifications**

Mediation analyses were conducted using cohort-specific models to account for different outcome types and available covariates. The mediation analysis was designed to explore the causal mechanism by which PTSS influences CVD through CpG methylation. Here, CpGs acted as mediators, and cell type proportions were included as covariates to control for potential confounding due to cellular heterogeneity in methylation profiles. This adjustment ensures that observed mediation effects are not biased by differences in cell composition, which is critical for accurate interpretation of epigenetic mediation. The discovery cohort (DNHS), we used logistic regression for the CVD outcome, and a linear regression model is used for the continuous variable, which is MRS_CVD.

**In DNHS:**

CVD ~ PTSS + CpG + Age Categories + Sex + Smoking Score + BMI Categories + Trauma number + Ancestry PCs (PC2 + PC3) + Cell Proportions (CD8T + CD4T + NK + Bcell + Mono)

For the replication cohorts using continuous MRS_CVD outcomes, we employed linear regression models for both GTP and NHS.

**In GTP:**

MRS_CVD ~ PTSS + CpG + Age Categories + Sex + Smoking Score + Trauma number + Ancestry PCs (PC2 + PC3) + Cell Proportions (CD8T + CD4T + NK + Bcell + Mono)

**In NHS:**

MRS_CVD ~ PTSS + CpG + Age Categories + Smoking Score + BMI Categories + Trauma number + Ancestry PCs (PC2 + PC3) + Cell Proportions (CD8T + CD4T + NK + Bcell + Mono)

All models were adjusted for the same covariates listed above, and statistical significance was determined based on 95% confidence intervals derived from 1000 bootstrap simulations.

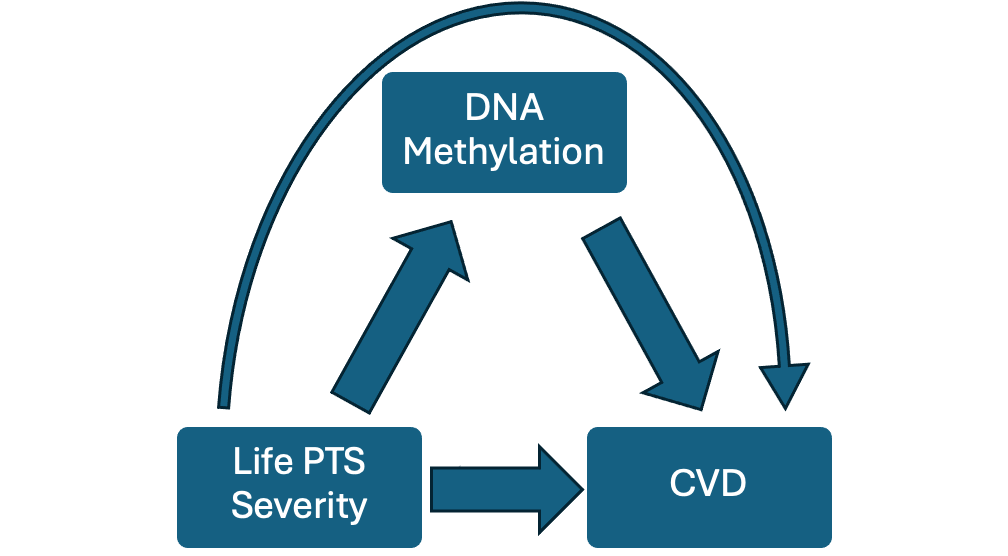

Figure 2. Visualization of Mediation Effect

CpG sites with significant mediation effects were retained for further biological interpretation, providing insight into the potential epigenetic mechanisms linking psychological stress and cardiovascular disease.

### Gene Ontology Enrichment Analysis

To further explore the functional relevance of our findings, we conducted gene ontology (GO) enrichment analyses using the missMethyl (v1.42.0) R package. **The** seven mediators CpG sites identified as potential mediators of the PTSS–CVD relationship were tested for GO pathway enrichment using a background set of 7,693 CpGs mapped to CoRSIVs. All enrichment results were adjusted for multiple testing using the Benjamini-Hochberg method, with a false discovery rate (FDR) of 5% used to determine significance.

### Code availability

The scripts generated to perform Cardiovascular Disease Risk Prediction Model, Methylation marker prediction associated with CVD, Mediation analysis, and Enrichments analysis are available in GitHub (<https://github.com/uddin-research-group-at-usf/CorSIVs_Project>).
